# Bacteremia and Blood Culture Utilization During COVID-19 Surge in New York City

**DOI:** 10.1101/2020.05.05.20080044

**Authors:** Jorge Sepulveda, Lars F. Westblade, Susan Whittier, Michael J. Satlin, William G. Greendyke, Justin G. Aaron, Jason Zucker, Donald Dietz, Magdalena Sobieszczyk, Justin J. Choi, Dakai Liu, Kelvin Espinal, Sarah Russell, Dennis Camp, Charles Connelly, Daniel A. Green

## Abstract

A surge of patients with coronavirus disease 2019 (COVID-19) presenting to New York City hospitals in March 2020 led to a sharp increase in the utilization of blood cultures, which overwhelmed the capacity of automated blood culture instruments. We sought to evaluate the utilization and diagnostic yield of blood cultures during the COVID-19 pandemic to determine prevalence and common etiologies of bacteremia, and to inform a diagnostic approach to relieve blood culture overutilization. We performed a retrospective cohort analysis of 88,201 blood cultures from 28,011 patients at a multicenter network of hospitals within New York City to evaluate order volume, positivity rate, time to positivity, and etiologies of positive cultures in COVID-19. Ordering volume increased by 34.8% in the second half of March 2020 compared to the first half of the month. The rate of bacteremia was significantly lower among COVID-19 patients (3.8%) than COVID-19 negative patients (8.0%) and those not tested (7.1%), p < 0.001. COVID-19 patients had a high proportion of organisms reflective of commensal skin microbiota, reducing the bacteremia rate to 1.6% when excluded. More than 98% of all positive cultures were detected within 4 days of incubation. Bloodstream infections are very rare for COVID-19 patients, which supports the judicious use of blood cultures in the absence of compelling evidence for bacterial coinfection. Clear communication with ordering providers is necessary to prevent overutilization of blood cultures during COVID-19 surges, and laboratories should consider shortening the incubation period from 5 days to 4 days to free additional capacity.

## Introduction

The rapid spread of severe acute respiratory syndrome coronavirus 2 (SARS-CoV-2) across New York City in March 2020 led to an unprecedented strain on hospital resources, including shortages of beds, ventilators, personal protective equipment, and diagnostic materials such as laboratory reagents and nasopharyngeal swabs ^1–3^. The surge of febrile patients to our network of hospitals in New York City led to a sudden and dramatic increase in the number of blood cultures received in our laboratories, which overwhelmed the capacity of our automated blood culture instruments.

While blood cultures are an essential tool for the diagnosis and management of bloodstream infections among patients presenting to the emergency department and among inpatients, data are lacking on their utility for patients with suspected or confirmed coronavirus disease 19 (COVID-19). While many patients with severe COVID-19 are treated with empiric antibiotics for potential bacterial co-infections, the rate of bacteremia among these patients is unknown, and the benefit of empiric antibiotic therapy is unproven. Frequent ordering of blood cultures for patients with COVID-19 may overwhelm a laboratory’s capacity to perform and result these tests, which may negatively impact the overall benefit of testing for the entire medical center. Therefore, we sought to evaluate both the utilization and diagnostic yield of blood cultures during a surge of COVID-19 patients presenting to our hospitals, including positivity rates for patients with and without COVID-19, as well as the most common causes of bacteremia among COVID-19 patients. We also relay strategies for diagnostic stewardship to mitigate challenges that may arise from a surge of blood culture orders.

## Methods

### Study Design

A retrospective cohort study was conducted on patients with blood cultures performed at NewYork-Presbyterian Hospitals located throughout New York City from January 1, 2020 to March 31, 2020. Corresponding data from January 1, 2019 to March 31, 2019 were collected to establish a seasonal historic baseline of blood culture ordering and positivity. Records were extracted from the laboratory information system (Cerner Millennium, Cerner, North Kansas City, MO) using a Cerner Command Language query and included information on performing facility, SARS-CoV-2 reverse transcription-polymerase chain reaction (RT-PCR) result, blood culture result, organism(s) identified, and blood culture collection date and time. After the study period concluded, the blood culture incubation period was reduced from 5 days to 4 days to free additional space on the instruments. In a subset of patients for whom data were available, the interval from time of blood culture collection to time of Gram stain was used to calculate the time to blood culture positivity during the study period, which was used to examine the predicted effect of this intervention.

### Laboratory Methods

Blood cultures were incubated on BACTEC FX (Becton, Dickinson and Co., Franklin Lakes, NJ) or VersaTrek (Thermo Fisher Scientific, Inc., Waltham, MA) instruments for a maximum of 5 days. SARS-CoV-2 RT-PCR testing was performed in-house with the following assays: cobas SARS-CoV-2 (Roche Molecular Systems, Inc., Branchburg, NJ), Xpert Xpress SARS-CoV-2 (Cepheid, Sunnyvale, CA), RealStar SARS-CoV-2 (Altona Diagnostics USA, Inc., Plain City, OH), and a laboratory-developed test developed by the Wadsworth Center at the New York State Department of Health.

### Participants

A total of 88,201 blood cultures from 28,011 patients were included from the following hospitals within the NewYork-Presbyterian network: Columbia University Irving Medical Center (32,788), Weill-Cornell Medical Center (26,794), Allen Hospital (6,053), Queens Hospital (16,913), and Lower Manhattan Hospital (5,653). Patients were stratified by SARS-CoV-2 RT-PCR result as positive, negative, or not tested. For the purposes of classifying blood cultures by SARS-CoV-2 RT-PCR status, we used the following criteria:

1. Blood cultures were labeled as SARS-CoV-2 status “Positive” if the blood culture was performed within 2 days of a positive SARS-CoV-2 RT-PCR result and for all subsequent blood cultures after a positive SARS-CoV-2 RT-PCR result.
2. Blood cultures were labeled as SARS-CoV-2 status “Negative” if the blood culture was performed within 2 days of a negative SARS-CoV-2 RT-PCR result and for all subsequent blood cultures, unless the patient had a subsequent positive SARS-CoV-2 RT-PCR result, for which the status was changed to “Positive” for any blood cultures performed within 2 days of the positive SARS-CoV-2 RT-PCR result.
3. All other blood cultures were labeled as SARS-CoV-2 status “Not tested.”

The 2-day interval was used to account for turnaround time from test ordering to SARS-CoV-2 test results, as blood culture and SARS-CoV-2 RT-PCR tests ordered on the same day may have taken up to 2 days for the SARS-CoV-2 RT-PCR result to become available.

This study was approved by the Institutional Review Boards of Columbia University Irving Medical Center and Weill-Cornell Medicine.

### Data Analysis

All data analysis was performed with the R statistical language version 3.6.3 *(R Core Team (2020). R: A language and environment for statistical computing. R Foundation for Statistical Computing, Vienna, Austria. URL https://www.R-project.org)*. Blood culture volumes and positivity rates were calculated and graphed by day, with a moving regression line estimated by the default *geom_smooth* function of the *ggplot2* R package version 3.3.0, namely locally estimated scatterplot smoothing (LOESS) for data with less than 1000 points and generalized additive model (GAM) for data with more than 1000 points. The regression lines were overlayed in the scatterplot to analyze directional trends.

Volumes and positivity rates were also stratified by SARS-CoV-2 RT-PCR result using the rules specified above, and by various patient categories. Bacterial and fungal etiologies of blood cultures were also collected and stratified by SARS-CoV-2 RT-PCR result. Differences in continuous data between groups were assessed by one-way analysis of variation (ANOVA), whereas categorical data were analyzed by Pearson’s chi-squared analysis. Categorical data were graphed by the *assoc* function of the *vcd* R package (version 1.4.7) and colored according to Pearson’s residuals to demonstrate sources of difference between observed and expected proportions in different groups.

## Results

### Blood Culture Volumes and Positivity Rate

During the study period, blood culture volumes rose substantially during the month of March 2020 (**Figure 1**). Overall, 8,784 blood cultures were performed during the second half of March 2020, representing a 34.8% increase from the first half of the month. Patients who were positive for SARS-CoV-2 accounted for the majority of the increased blood culture ordering. Notably, the increased ordering among COVID-19 patients was not primarily attributable to repeated ordering, as 48.2% of COVID-19 patients had more than 2 blood culture sets drawn, compared to 66.2% of COVID-19 negative patients and 62.1% of patients not tested for SARS-CoV-2 (p <0.001, **Supplemental Tables 1 and 2**)

**Figure 1:**
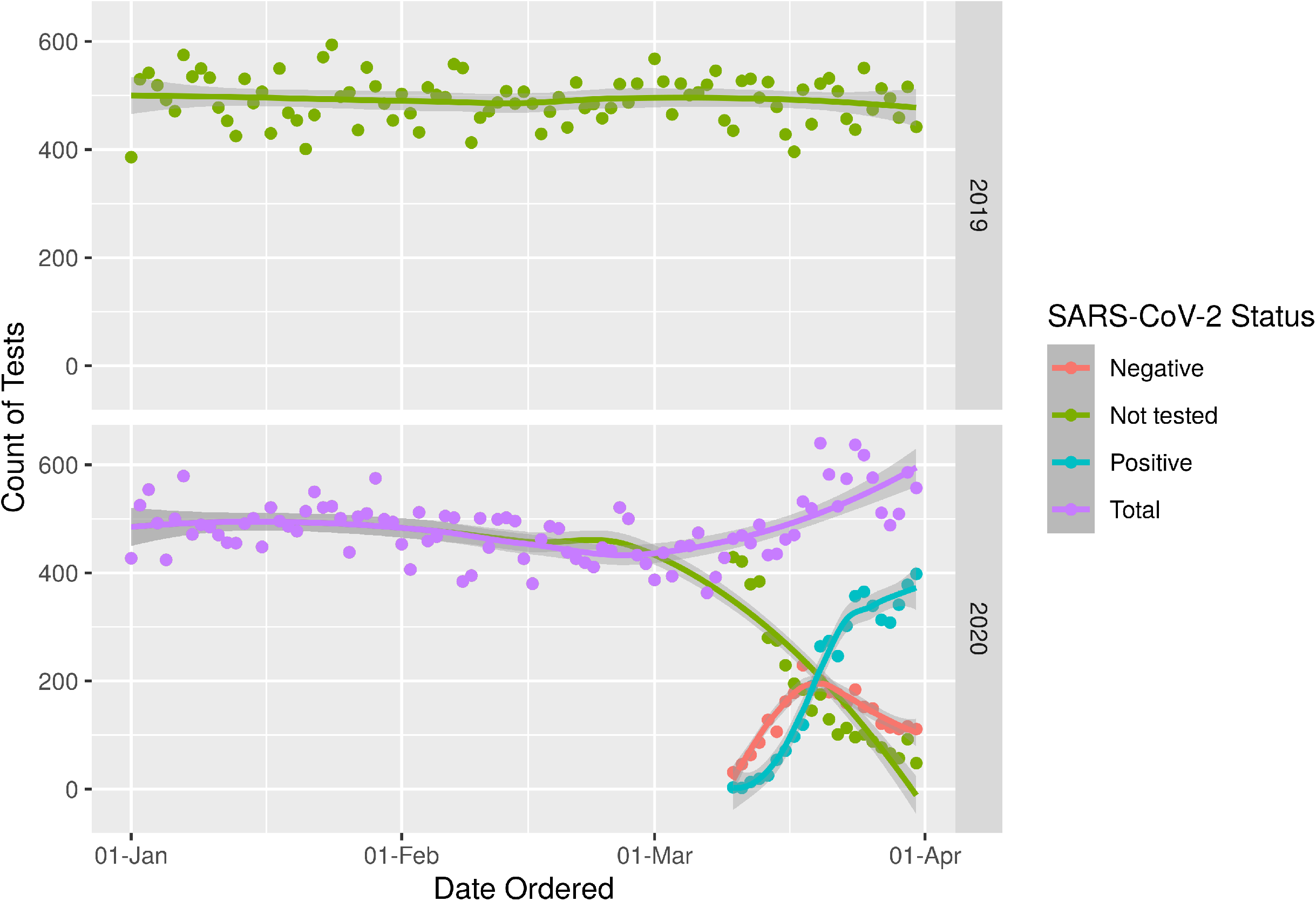
Blood culture ordering volume by day in 2019 and 2020. Total blood cultures are shown in purple and are broken down by SARS-CoV-2 status of “Positive” in blue, “Negative” in red, and “Not tested” in green.

**Table 1A.** Distribution of blood culture results by SARS-CoV-2 status

| **SARS-CoV-2 Status** | Jan-Feb, 2019<br>(N=29019) | Mar 1-15, 2019<br>(N=7601) | Mar 16-31, 2019<br>(N=7688) | Jan-Feb, 2020<br>(N=28593) | Mar 1-15, 2020<br>(N=6518) | Mar 16-31, 2020<br>(N=8784) | Total<br>(N=88203) |
| --- | --- | --- | --- | --- | --- | --- | --- |
| **Blood Culture Result** |  |  |  |  |  |  |  |
| **Not tested** |  |  |  |  |  |  |  |
| Negative | 27022<br>(93.1%) | 7074<br>(93.1%) | 7065<br>(91.9%) | 26520<br>(92.7%) | 5556<br>(93.5%) | 1779<br>(93.8%) | 75016<br>(92.9%) |
| Positive | 1997<br>(6.9%) | 527<br>(6.9%) | 623<br>(8.1%) | 2073<br>(7.3%) | 386<br>(6.5%) | 117<br>(6.2%) | 5723<br>(7.1%) |
| **Negative** |  |  |  |  |  |  |  |
| Negative | 0 | 0 | 0 | 0 | 431<br>(93.7%) | 2317<br>(91.7%) | 2748<br>(92.0%) |
| Positive | 0 | 0 | 0 | 0 | 29 (6.3%) | 209<br>(8.3%) | 238<br>(8.0%) |
| **Positive** |  |  |  |  |  |  |  |
| Negative | 0 | 0 | 0 | 0 | 110<br>(94.8%) | 4198<br>(96.2%) | 4308<br>(96.2%) |
| Positive | 0 | 0 | 0 | 0 | 6 (5.2%) | 164<br>(3.8%) | 170<br>(3.8%) |

**Table 1B.**
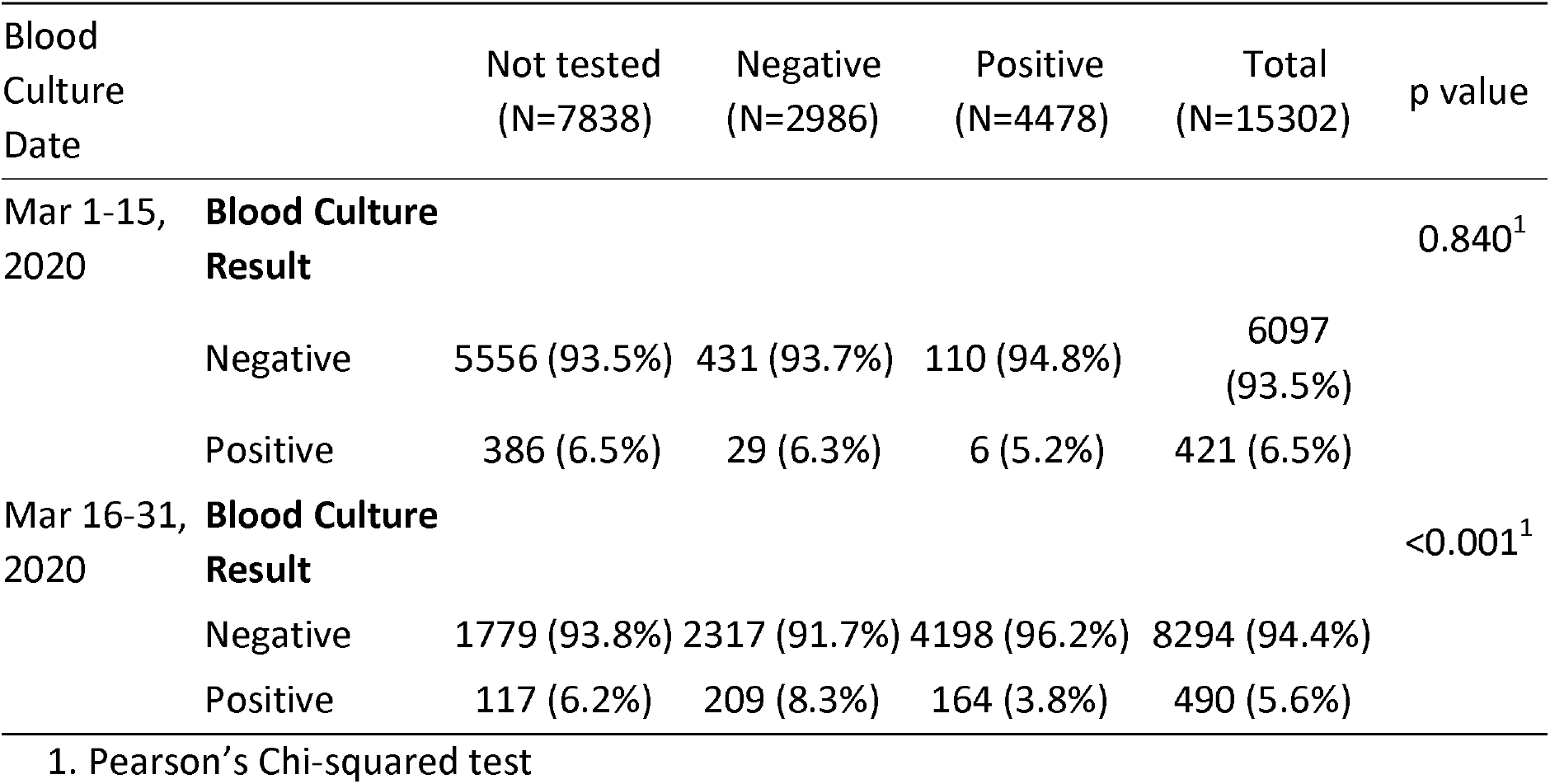
Distribution of blood culture results collected in March 2020 by SARS-CoV-2 status.

| Blood Culture Date | Not tested<br>(N=7838) | Negative<br>(N=2986) | Positive<br>(N=4478) | Total<br>(N=15302) | p value |
| --- | --- | --- | --- | --- | --- |
| Mar 1-15, 2020 |  |  |  |  | 0.840<sup>1</sup> |
| **Blood Culture Result** |  |  |  |  |  |
| Negative | 5556 (93.5%) | 431 (93.7%) | 110 (94.8%) | 6097<br>(93.5%) |  |
| Positive | 386 (6.5%) | 29 (6.3%) | 6 (5.2%) | 421 (6.5%) |  |
| Mar 16-31, 2020 |  |  |  |  | <0.001<sup>1</sup> |
| **Blood Culture Result** |  |  |  |  |  |
| Negative | 1779 (93.8%) | 2317 (91.7%) | 4198 (96.2%) | 8294 (94.4%) |  |
| Positive | 117 (6.2%) | 209 (8.3%) | 164 (3.8%) | 490 (5.6%) |  |

1. Pearson's Chi-squared test

The blood culture positivity rate was significantly lower for patients that tested positive for SARS-CoV-2 (3.8%) than for patients that tested negative for SARS-CoV-2 (8.0%) or for patients that were not tested (7.1%, p < 0.001, **Table 1**). As additional COVID-19 patients presented to the hospital throughout the month of March 2020, the overall rate of blood culture positivity continued to decrease further (**Figure 2**).

**Figure 2:**
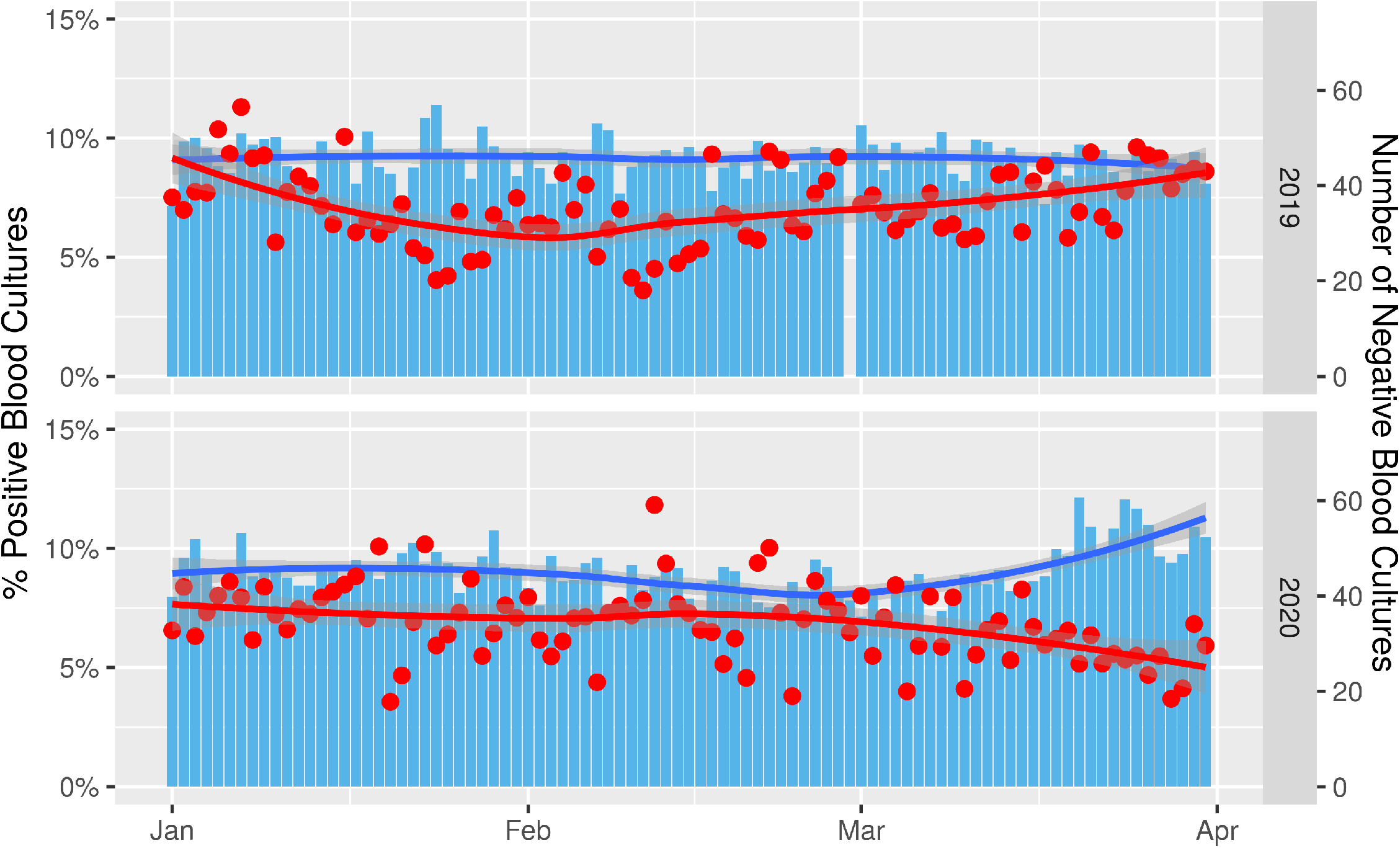
Rate of blood culture positivity by day in 2019 and 2020. The percentage of positive blood cultures is shown in red and the total number of negative blood cultures is shown in blue.

### Etiologies of Bacteremia

Among patients with positive blood cultures, COVID-19 patients had a significantly higher proportion of cultures that likely represented contamination with normal skin microbiota than all other groups (**Figure 3 and Table 2**). Organisms were labeled as likely contaminants if they were isolated only once per patient and belonged to groups generally defined as commensal skin microbiota.^4^ Coagulase-negative *Staphylococcus* species accounted for 59.7% of all positive cultures among COVID-19 patients, compared to 32.0% among patients that tested negative for SARS-CoV-2, and 29.8% among patients that were not tested for SARS-CoV-2 in 2020 (p < 0.001). *Corynebacterium* species, *Bacillus* species, and *Micrococcus* species were also seen more frequently among COVID-19 patients (**Supplemental Table 3**). When potential contaminants were excluded, the rate of bacteremia for COVID-19 patients decreased to 1.6%, which was significantly lower than the rate of bacteremia excluding contaminants among COVID-19 negative patients (5.9%) and during the same period in 2019 (5.7%, p < 0.001, **Table 3**). The most common causes of true bacteremia among COVID-19 patients were *Escherichia coli* (16.7%), *Staphylococcus aureus* (13.3%), *Klebsiella pneumoniae* (10.0%), and *Enterobacter cloacae* complex (8.3%) (**Supplemental Table 3**). None of these pathogens were overrepresented among COVID-19 patients compared to the other groups.

**Figure 3:**
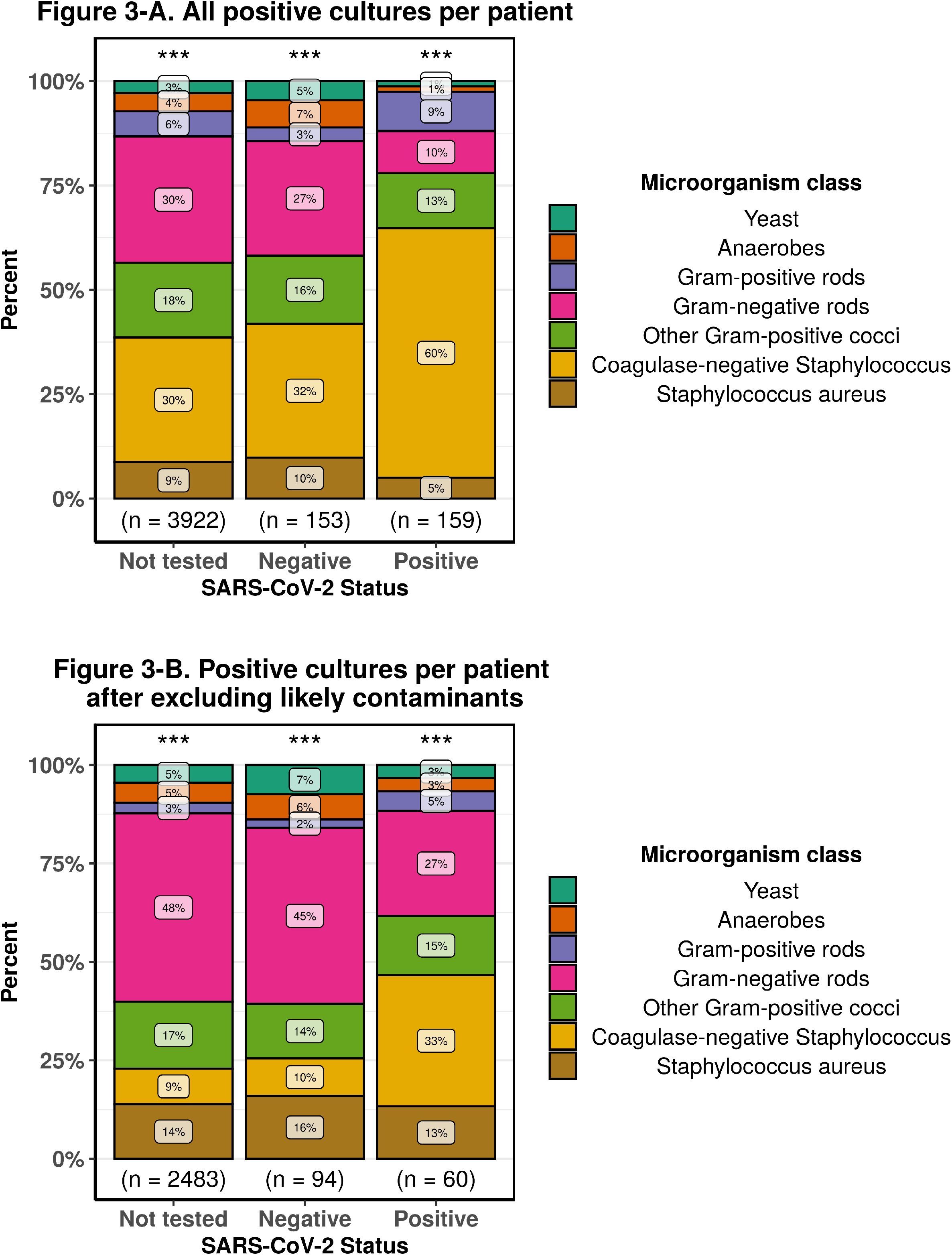
Frequency of microorganisms identified from positive blood cultures stratified by SARS-CoV-2 status. Each microorganism was counted once per patient and grouped by the categories specified in the figure legends. A. All microorganisms isolated were counted. B. Likely skin contaminant were excluded. *** Pearson’s chi-square p-value < 0.001.

**Table 2.**
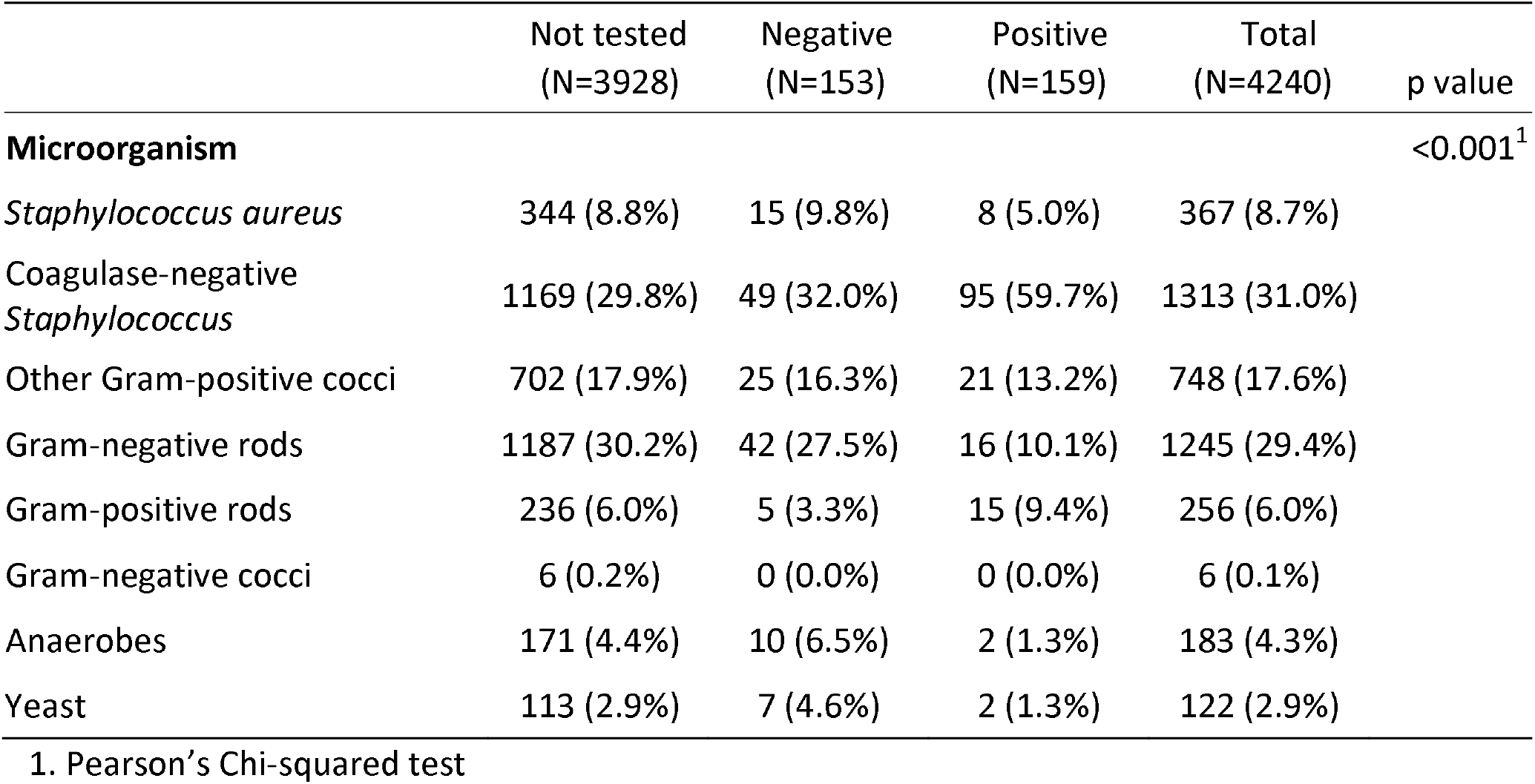
Most frequent microorganism groups isolated from blood cultures by SARS-CoV-2 status. Each species was counted once per patient, independently of how often it was isolated from each patient, then grouped by the conventional categories.

**Table 3.**
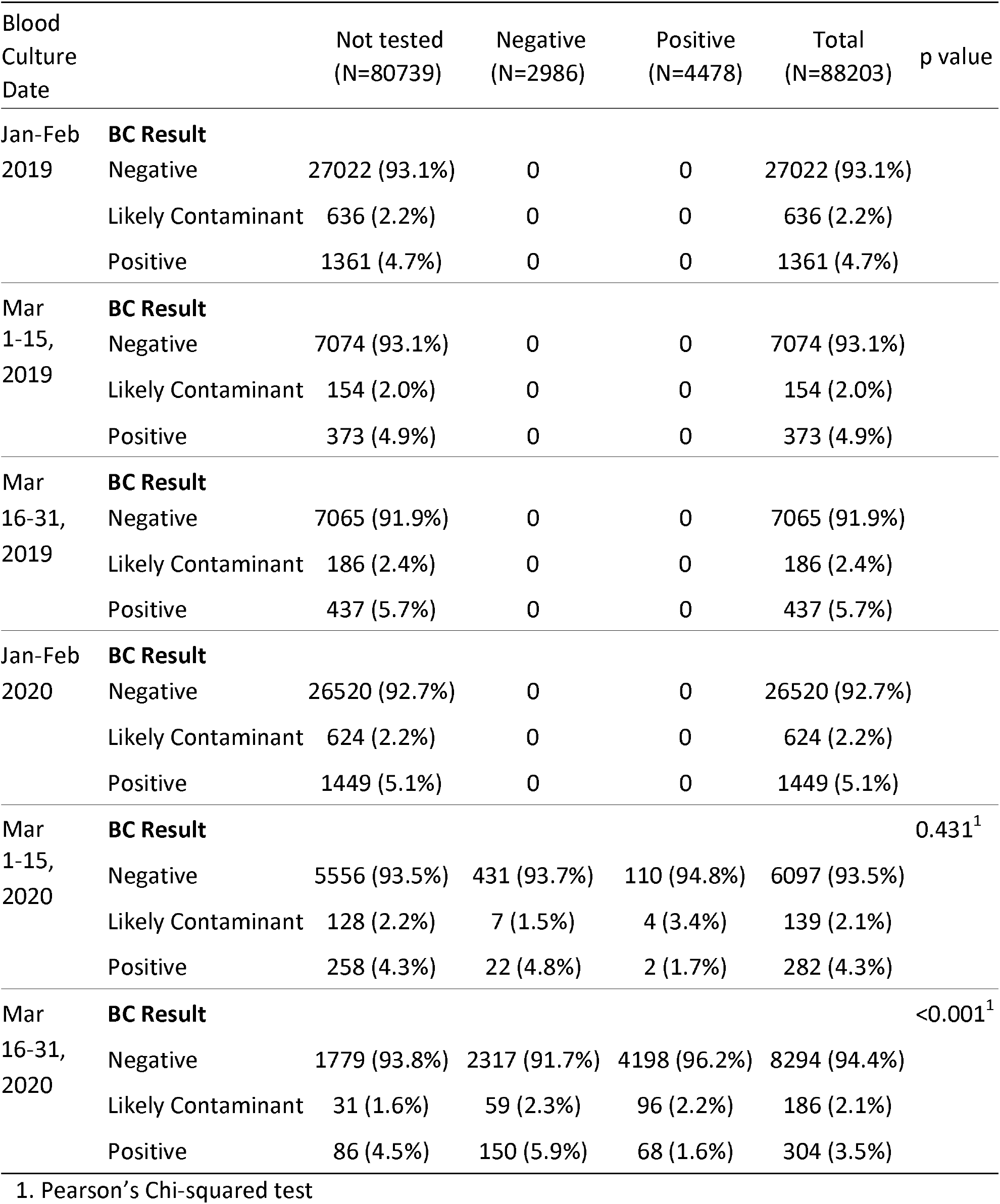
Distribution of blood culture results by date and SARS-CoV-2 Status. Organisms were labeled as likely contaminants if they were isolated only once per patient and belong to the groups generally defined as skin contaminants

| Blood Culture Date |  | Not tested<br>(N=80739) | Negative<br>(N=2986) | Positive<br>(N=4478) | Total<br>(N=88203) | p value |
| --- | --- | --- | --- | --- | --- | --- |
| Jan-Feb 2019 | **BC Result** |  |  |  |  |  |
|  | Negative | 27022 (93.1%) | 0 | 0 | 27022 (93.1%) |  |
|  | Likely Contaminant | 636 (2.2%) | 0 | 0 | 636 (2.2%) |  |
|  | Positive | 1361 (4.7%) | 0 | 0 | 1361 (4.7%) |  |
| Mar 1-15, 2019 | **BC Result** |  |  |  |  |  |
|  | Negative | 7074 (93.1%) | 0 | 0 | 7074 (93.1%) |  |
|  | Likely Contaminant | 154 (2.0%) | 0 | 0 | 154 (2.0%) |  |
|  | Positive | 373 (4.9%) | 0 | 0 | 373 (4.9%) |  |
| Mar 16-31, 2019 | **BC Result** |  |  |  |  |  |
|  | Negative | 7065 (91.9%) | 0 | 0 | 7065 (91.9%) |  |
|  | Likely Contaminant | 186 (2.4%) | 0 | 0 | 186 (2.4%) |  |
|  | Positive | 437 (5.7%) | 0 | 0 | 437 (5.7%) |  |
| Jan-Feb 2020 | **BC Result** |  |  |  |  |  |
|  | Negative | 26520 (92.7%) | 0 | 0 | 26520 (92.7%) |  |
|  | Likely Contaminant | 624 (2.2%) | 0 | 0 | 624 (2.2%) |  |
|  | Positive | 1449 (5.1%) | 0 | 0 | 1449 (5.1%) |  |
| Mar 1-15, 2020 | **BC Result** |  |  |  |  | 0.431<sup>1</sup> |
|  | Negative | 5556 (93.5%) | 431 (93.7%) | 110 (94.8%) | 6097 (93.5%) |  |
|  | Likely Contaminant | 128 (2.2%) | 7 (1.5%) | 4 (3.4%) | 139 (2.1%) |  |
|  | Positive | 258 (4.3%) | 22 (4.8%) | 2 (1.7%) | 282 (4.3%) |  |
| Mar 16-31, 2020 | **BC Result** |  |  |  |  | <0.001<sup>1</sup> |
|  | Negative | 1779 (93.8%) | 2317 (91.7%) | 4198 (96.2%) | 8294 (94.4%) |  |
|  | Likely Contaminant | 31 (1.6%) | 59 (2.3%) | 96 (2.2%) | 186 (2.1%) |  |
|  | Positive | 86 (4.5%) | 150 (5.9%) | 68 (1.6%) | 304 (3.5%) |  |

1. Pearson's Chi-squared test

### Incubation Period

Among the subset of 1859 positive blood cultures for which incubation period could be reliably assessed, the vast majority (88.2%) signaled positive within 1-2 days of incubation, with an additional 7.0% signaling positive on day 3 and 3.0% signaling positive on day 4 (**Figure 4 and Supplemental Table 4**). Only 1.8% of all blood cultures signaled positive on day 5, many of which yielded normal skin microbiota (**Supplemental Table 5**). Among COVID-19 patients, 97.3% of positive cultures signaled positive within 3 days of incubation, with one culture positive on the 4^th^ day for *Cutibacterium acnes* and one culture positive on the 5^th^ day for *Candida albicans*.

**Figure 4:**
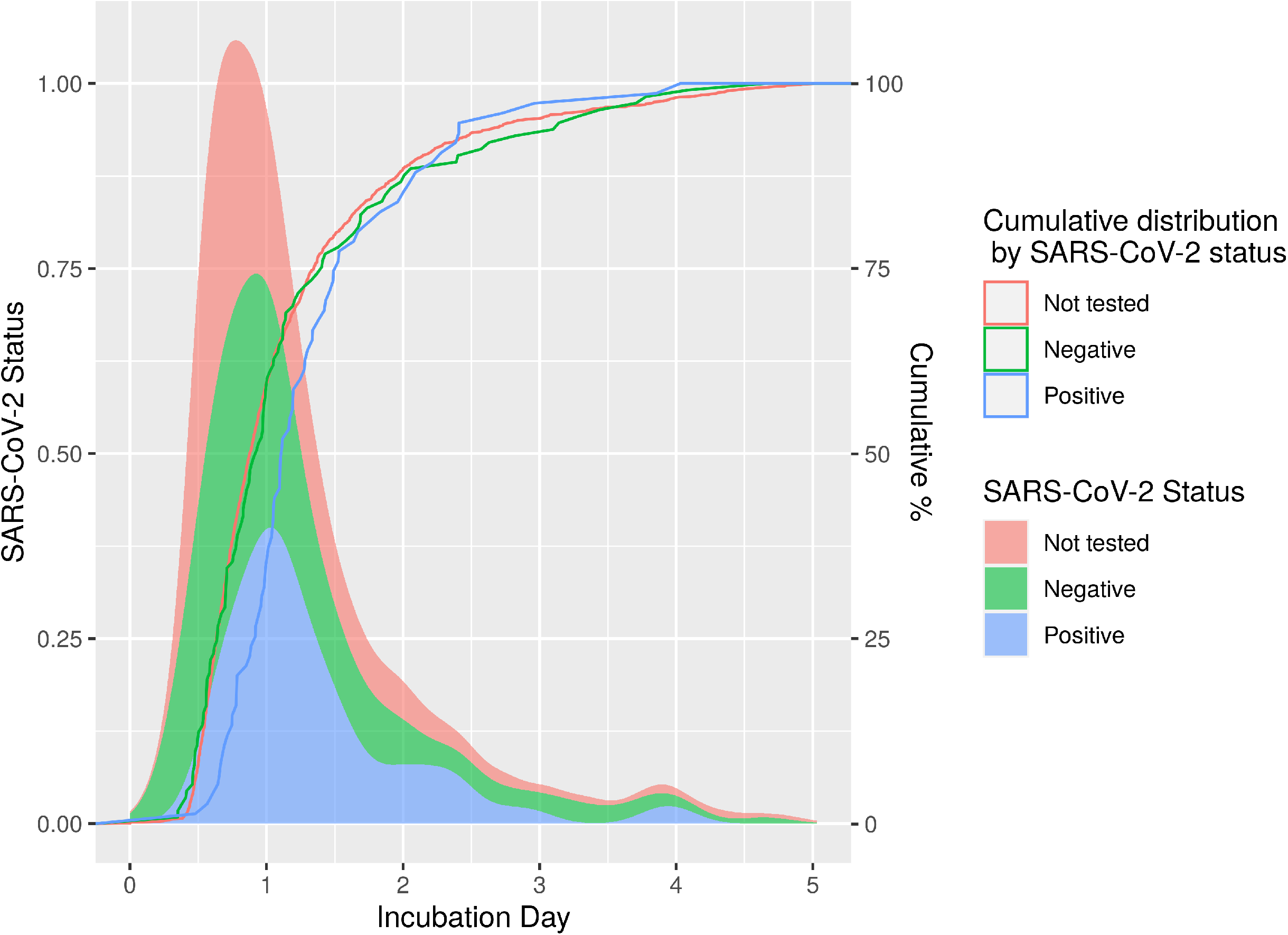
Time to positivity of positive blood cultures stratified by SARS-CoV-2 status. Time to positivity was calculated from the time of collection to the first positive signal as recorded by the gram stain date/time. The scaled density distribution of the time to positivity is shown as color-filled density plots and the cumulative distribution of positivity over time is represented by solid lines.

## Discussion

Beginning in March 2020, a surge of COVID-19 patients presenting to our network of hospitals in New York City, the current epicenter of the global COVID-19 pandemic^5^, led to a dramatic increase in the utilization of blood cultures. In patients presenting with severe febrile illness, blood cultures are essential in ruling out bacterial infection and guiding appropriate antibiotic utilization. However, we found a very low rate of bacteremia among patients diagnosed with COVID-19, implying a remarkably low diagnostic yield of blood cultures for COVID-19 patients. When excluding likely contaminants, COVID-19 patients had bacteremia rates that were less than one third of the baseline rate from 2019. These data demonstrate that bloodstream infections appear to be very rare for COVID-19 patients, and suggest that empiric antibiotics may not be useful in the absence of compelling evidence of an accompanying bacterial infection. Notably, we did not evaluate other bacterial infections such as bacterial pneumonia, although other studies have shown low levels of procalcitonin among COVID-19 patients, arguing that bacterial superinfection may be uncommon ^6–8^.

As medical centers across the United States prepare for anticipated waves of COVID-19 patients, our data may be used to justify the judicious utilization of blood cultures to preserve the operational capacity of diagnostic laboratories and to promote antimicrobial stewardship efforts to reduce unnecessary antibiotic administration. Overordering of blood cultures during a COVID-19 surge within our hospital network resulted in culture volumes that exceeded the capacity of our automated instruments, requiring additional staff to manually process these cultures at a time when staffing and supplies were already constrained. Our experience should serve as a caution to other medical centers that overordering of blood cultures during patient surges can overwhelm laboratory capacity and may negatively impact the quality of results for all patients.

As a harm-reduction measure, we decreased the incubation period of blood cultures from 5 days to 4 days after the study period concluded, which freed additional space to allow for timely processing of incoming cultures. Our data demonstrate that this intervention likely had little to no adverse effect on patient care, as all but one COVID-19 patients with positive cultures during the study period signaled positive within 4 days, and only 1.8% of all positive cultures signaled positive on the 5 ^th^ day, many of which were positive for normal skin microbiota. Previous studies have also shown that decreasing the incubation of blood cultures to 4 days ^9^ or even 3 days ^10,11^ has minimal effect on positivity rates, particularly for clinically significant bacteria.

To our knowledge, this study is the first to examine blood culture utilization among COVID-19 patients. The inclusion of over 88,000 patient cultures is a major strength of the study design, as is the multicenter analysis from a wide geographic catchment area in New York City, which increases the generalizability of our results. Limitations of the study include paucity of data on other bacterial co-infections and lack of data on patient antibiotic utilization to demonstrate how blood culture utilization impacted therapy.

In summary, we observed an overutilization of blood cultures during a surge of COVID-19 patients to our network of medical centers in New York City and found a very low rate of bloodstream infections among COVID-19 patients. This overutilization was mitigated through a 4 day incubation with likely minimal impact on patient care. Clear communication with ordering providers and hospital leadership regarding the low yield of blood cultures is a necessary step to mitigate overordering and to preserve laboratory functionality during these periods. Laboratories should also consider reducing the incubation period of blood cultures from 5 days to 4 days to further increase their capacity.

## Data Availability

The data that support the findings of this study are available from the corresponding author, DG, upon reasonable request.

## Acknowledgments

*Potential conflicts of interest:* J.J.C. has received research support from Roche Diagnostics. All other authors have no conflicts.

